# Targeted BRCA1/BRCA2 Sequencing in a Bangladeshi Clinically Referred Cohort Identifies Candidate BRCA1 Loss-of-Function Variants and a Multi-Exon Deletion-Like CNV Signal

**DOI:** 10.64898/2026.05.11.26352643

**Authors:** Syed Muktadir Al Sium, Tanjina Akhtar Banu, Barna Goswami, Showti Raheel Naser, Md Ahashan Habib, Shahina Akter, Mst. Hosne Ara, Sheikh Md Selim Al Din, Ali Nafisa, Maksudur Rahman Nayem, Mohammad Fazle Alam Rabbi, Md Murshed Hasan Sarkar, Md Salim Khan

## Abstract

**Background:** Population-relevant BRCA1/BRCA2 data from Bangladesh are scarce, creating challenges for hereditary breast and ovarian cancer variant interpretation, counseling, and follow-up testing. We examined a clinically referred Bangladeshi cohort to characterize assay-derived BRCA1/BRCA2 short variants, sequencing-depth performance, and copy-number findings in a conservative pilot framework.

**Methods:** Twenty-three de-identified blood-derived DNA samples were assessed using a targeted BRCA1/BRCA2 next-generation sequencing workflow. Downstream analysis used assay-generated short-variant, coverage, and CNV outputs, with coordinates reported on hg19/GRCh37. Short variants were evaluated from high-confidence PASS/VCC-H calls, and CNV review incorporated both target-region and amplicon-level copy-number patterns.

**Results:** After removal of four low-VAF review observations, the primary germline-compatible dataset comprised 304 short-variant observations representing 34 unique variants. Both BRCA1 and BRCA2 contributed comparable variant burdens, while the overall profile was mainly composed of missense and synonymous changes. Six sample-specific heterozygous BRCA1 truncating candidates were observed, including five frameshift variants and one stop-gain variant. Protein-level mapping placed these events across the central-to-C-terminal portion of BRCA1. Sequencing depth was consistently high across the targeted regions, with all 4,255 amplicon-sample measurements exceeding 280x and 99.91% reaching at least 500x. Copy-number analysis highlighted one candidate BRCA1 multi-exon deletion-like event involving exons 15-20 in BCSIR-BRCA-21, with unresolved partial exon 14 involvement.

**Conclusions:** This study provides an initial Bangladesh-focused targeted BRCA1/BRCA2 dataset and identifies candidate short-variant and CNV findings for validation. These findings should be interpreted as analytical candidates only and require confirmatory testing and expert clinical curation before any clinical application. The cohort is referral-enriched and should not be used to infer population prevalence.

## Introduction

Breast cancer is a major global cancer burden, and inherited susceptibility contributes to early-onset disease, familial clustering, contralateral breast cancer risk, and ovarian cancer risk (Ferlay et al., 2021; Kuchenbaecker et al., 2017). Pathogenic germline variants in BRCA1 and BRCA2 compromise homologous recombination repair and have direct implications for genetic counseling, cascade testing, risk-reducing strategies, surveillance, and treatment selection (Kuchenbaecker et al., 2017; Richards et al., 2015).

Variant interpretation remains uneven across populations. Public repositories such as ClinVar have improved access to submitted variant interpretations, but underrepresented populations continue to face disproportionate uncertainty because of limited ancestry-matched frequency data, incomplete segregation evidence, and limited local curation resources (Landrum et al., 2018). South Asian studies have shown substantial heterogeneity in reported BRCA1/BRCA2 variants across countries and communities, emphasizing that regional findings cannot be assumed to transfer directly between neighboring populations (Kharel et al., 2022).

Bangladesh-specific BRCA1/BRCA2 data remain scarce. Earlier Bangladeshi studies have often focused on selected exons or specific variants rather than assay-wide targeted NGS profiles, leaving a gap in locally relevant variant interpretation resources (Chowdhury et al., 2020; Nishat et al., 2019). For settings with limited hereditary cancer testing infrastructure, early assay-wide datasets are useful not because they establish prevalence, but because they identify candidate variants for confirmation, highlight workflow challenges, and support development of local genetic counseling and confirmatory testing pathways.

Here, we describe BRCA1/BRCA2 short variants, coverage-based sequencing QC, and CNV signals identified in a Bangladeshi clinically referred cohort using a targeted BRCA1/BRCA2 NGS workflow. The analysis was restricted to assay-derived short-variant and CNV outputs, with embedded database versions reported explicitly. We distinguish shared polymorphic signals from candidate analytically relevant findings and avoid clinical overinterpretation in the absence of orthogonal confirmation.

## Materials and Methods

### Study design, participants, clinical indication, and ethics

This was a pilot analytical study of 23 de-identified blood-derived DNA samples designated BCSIR-BRCA-01-06 and BCSIR-BRCA-08-24, processed at the Genomic Research Laboratory, BCSIR, Bangladesh. The cohort was referral-enriched and was not designed to estimate BRCA1/BRCA2 variant prevalence in the Bangladeshi population.

The cohort primarily comprised clinically referred individuals evaluated for suspected breast cancer or related clinical concern on the basis of referral documentation and laboratory intake records. One participant was unaffected at presentation and underwent predictive screening because of a documented family history. Because the study was retrospective and referral-based, participants were not enrolled using uniform hereditary cancer testing criteria. Available clinical and referral metadata were abstracted from laboratory intake forms and accompanying referral documents, including age, referral indication, family-history status, selected clinical descriptors where available, and TNBC status when explicitly documented. Referral indication was categorized descriptively as suspected/affected breast cancer-related referral, family-history-based predictive screening, or incompletely documented referral indication. Because these data were incomplete and non-uniformly recorded, clinical variables were used only for descriptive cohort characterization and not for formal genotype–phenotype analysis.

This study was conducted in accordance with the national laws and regulations of Bangladesh and the WMA Declaration of Helsinki for medical research involving human subjects. Ethical approval was obtained from the Ethical Committee of the National Institute of Cancer Research and Hospital (NICRH), Mohakhali, Dhaka-1212, Bangladesh (Ref. No. NICRH/Ethics/2019/525; approval date: 22 September 2019). Written informed consent was obtained from all participants before inclusion in the study.

### Library preparation, sequencing, and AmpliconSuite analysis

Library preparation was performed using a Devyser targeted BRCA1/BRCA2 next-generation sequencing assay according to the manufacturer’s instructions (Capone et al., 2018). The assay targets BRCA1 and BRCA2 coding regions and assay-defined splice-adjacent regions. Sequencing was performed on an Illumina NextSeq 550 platform.

FASTQ outputs were analyzed using AmpliconSuite v3.8.1 with the Devyser_BRCA v1.1.1 assay-specific pipeline. Exported VCF, SNV, target-region CNV, amplicon-level CNV, and amplicon-level coverage files were used for downstream analysis. Variant coordinates were reported on hg19/GRCh37 according to the assay output. Transcript annotations were taken directly from exported VCF/SNV files, including BRCA1 NM_007294.4 and BRCA2 NM_000059.3. Variant nomenclature followed HGVS recommendations where applicable, and VCF handling followed standard VCF conventions (Danecek et al., 2011; den Dunnen et al., 2016).

### Coverage-based sequencing QC

Coverage-based QC was summarized from an amplicon-level coverage table containing 185 assay amplicons across the 23 samples. For each sample, minimum depth, median depth, mean depth, maximum depth, and the proportion of amplicons meeting coverage thresholds of 100x, 200x, 500x, and 1000x were calculated. These metrics were used to support analytical coverage adequacy for the targeted assay output. The coverage table was not treated as a complete substitute for full sequencing-run QC because it does not independently assess all metrics relevant to NGS performance.

### Short-variant filtering and analytical dataset

The Devyser VCF exports used the following filters: minimum variant allele fraction of 20%, mean variant quality score >=20, variant support by at least 10 reads, and restriction to assay-defined target regions. Only variants with FILTER = PASS and high variant-calling confidence (VCC=H) were considered for cohort-level analysis.

For the main germline-compatible analytical dataset, variants were retained if allele fraction and genotype were consistent with expected germline heterozygous or homozygous states. Low-VAF variants near the 20% filtering threshold, particularly those not assigned a heterozygous genotype, were retained only as review variants, excluded from the main germline-compatible summary figures, and not interpreted as confirmed germline findings.

### Annotation strategy and public database review

Variant annotations were extracted from the assay-derived VCF/SNV outputs. The embedded annotation database versions reported in the output were ClinVar_20190715 and dbSNP_b151. These annotations were used for descriptive summarization, while candidate analytically relevant variants were additionally checked against public database records during manuscript preparation.

For ClinVar summary plots, each variant observation was assigned to a single simplified category. Benign and likely benign labels were grouped as benign/likely benign; pathogenic and likely pathogenic labels were grouped as pathogenic/likely pathogenic; and records containing mixed benign, pathogenic, uncertain, or otherwise inconsistent labels were categorized as conflicting interpretations.

Candidate predicted loss-of-function variants were additionally checked against public database records, primarily ClinVar and the ClinGen Allele Registry, to contextualize the embedded assay annotations (Landrum et al., 2018; Pawliczek et al., 2018). This cross-check did not alter the primary callset, replace orthogonal confirmation, or constitute formal ACMG/AMP or ENIGMA-aligned clinical classification.

### Candidate BRCA1 loss-of-function analytical findings and IGV review

Candidate BRCA1 loss-of-function analytical findings were defined as Devyser PASS/VCC-H variants with germline-compatible allele fractions and predicted truncating consequences, including frameshift and stop-gain variants. Candidate variants were visually reviewed in IGV using available BAM/BAI files. Because orthogonal confirmation was not performed, these variants are reported as analytical candidates requiring confirmatory testing and expert clinical curation before any clinical reporting or clinical use.

### CNV analysis

CNV signals were evaluated using both target-region and amplicon-level CNV outputs from Devyser AmpliconSuite. Target-region CNV values were used for exon-level summarization, whereas amplicon-level CNV values were reviewed to determine whether candidate CNV signals were supported by multiple consecutive amplicons rather than isolated amplicon fluctuations. A candidate deletion-like CNV was considered when consecutive target-region values clustered around approximately 0.5 and were supported by concordant reductions across multiple amplicons. Candidate CNV events were visually reviewed in IGV where possible. Assay-derived CNV evidence was considered screening-level unless confirmed by MLPA, ddPCR, or another validated CNV confirmation method.

### Statistical analysis and visualization

Counts are presented as variant observations unless otherwise specified. Unique variants were defined using Devyser-derived genomic position, reference allele, alternate allele, gene, and transcript-level annotation. Descriptive statistics and visualizations were generated in R. Figures included gene-wise variant observation and unique-variant counts, consequence-class summaries, per-sample consequence distributions, protein-level lollipop plots, an oncoprint-style heatmap, a Devyser embedded ClinVar summary, and target-region/amplicon-level CNV plots. Because the SNV annotation output included multiple RefSeq transcript annotations for some BRCA1 variants, all variant-level analyses and HGVS reporting were standardized to the primary coding transcripts BRCA1 NM_007294.4 and BRCA2 NM_000059.3. Alternative BRCA1 transcript annotations were retained in raw exported files but were not counted as separate variant observations.

## Results

### Clinical metadata and referral context

The analytical cohort included 23 de-identified blood-derived DNA samples from clinically referred Bangladeshi individuals. Available metadata supported the referral-enriched nature of the cohort (**Table 1**). Twenty-two participants were referred in the context of suspected breast cancer or related clinical concern, while one unaffected participant underwent predictive screening because of a documented family history. Age data were available for 17 participants, with a median age of 36 years and a range of 23–64 years. Family-history information was documented for 15 participants, of whom 12 reported a positive family history of breast, ovarian, or related cancer. TNBC status was documented for only three participants, of whom two were reported as TNBC. Because clinical metadata were incomplete and were curated retrospectively from referral documentation and laboratory intake records, these variables were summarized descriptively and were not used for formal genotype–phenotype association testing.

**Table 1.** Available baseline clinical characteristics of the Bangladeshi BRCA referral cohort.

| Characteristic | Value |
| --- | --- |
| Total analytical cohort size | 23 |
| Age available, n | 17 |
| Age, median (range), years | 36 (23-64) |
| Suspected breast cancer / clinically referred, n | 22 |
| Family-history-based screening, n | 1 |
| Family history documented, n | 15 |
| Family history present, n | 12 |
| Family history absent among documented, n | 3 |

*Note: Clinical metadata were incomplete and were summarized descriptively only.*

### Coverage-based sequencing QC

The amplicon-level coverage table contained 185 amplicons across 23 samples, corresponding to 4,255 amplicon-sample measurements (Table 2). Overall mean depth was 2722.5x, median depth was 2141.0x, and the observed range was 284-15796x. All amplicon-sample measurements were at or above 100x and 200x. In addition, 99.91% of measurements were at or above 500x and 93.98% were at or above 1000x. Per-sample mean depth ranged from 1989.3x to 4823.6x, and per-sample median depth ranged from 1572.0x to 3980.0x. These coverage metrics supported inclusion of all 23 samples in the analytical cohort, while remaining limited to coverage-based QC rather than comprehensive run-level QC.

**Table 2.** Coverage-based sequencing QC summary.

| Metric | Value |
| --- | --- |
| Samples assessed | 23 |
| Amplicons assessed per sample | 185 |
| Total amplicon-sample measurements | 4,255 |
| Overall mean depth | 2722.5x |
| Overall median depth | 2141.0x |
| Overall depth range | 284-15796x |
| Measurements $\geq 100x$ | 100.0% |
| Measurements $\geq 200x$ | 100.0% |
| Measurements $\geq 500x$ | 99.91% |
| Measurements $\geq 1000x$ | 93.98% |
| Per-sample mean depth range | 1989.3-4823.6x |
| Per-sample median depth range | 1572.0-3980.0x |
| Values below 500x | 4 measurements; none below 100x or 200x |

### Short-variant callset

The assay-specific short-variant outputs contained 308 PASS high-confidence short-variant observations corresponding to 35 unique variants across the 23-sample analytical cohort. Four observations corresponded to a low-VAF BRCA2 c.68-7del splice-region/intronic deletion near the filtering threshold and were retained only as review variants. After excluding these low-VAF review observations from the main germline-compatible analytical set, 304 short-variant observations corresponding to 34 unique variants remained.

BRCA1 and BRCA2 contributed similar numbers of high-confidence short-variant observations and unique variants. BRCA1 accounted for 150 observations and 17 unique variants, while BRCA2 accounted for 154 observations and 17 unique variants (**Figure 1**).

**Figure 1.**
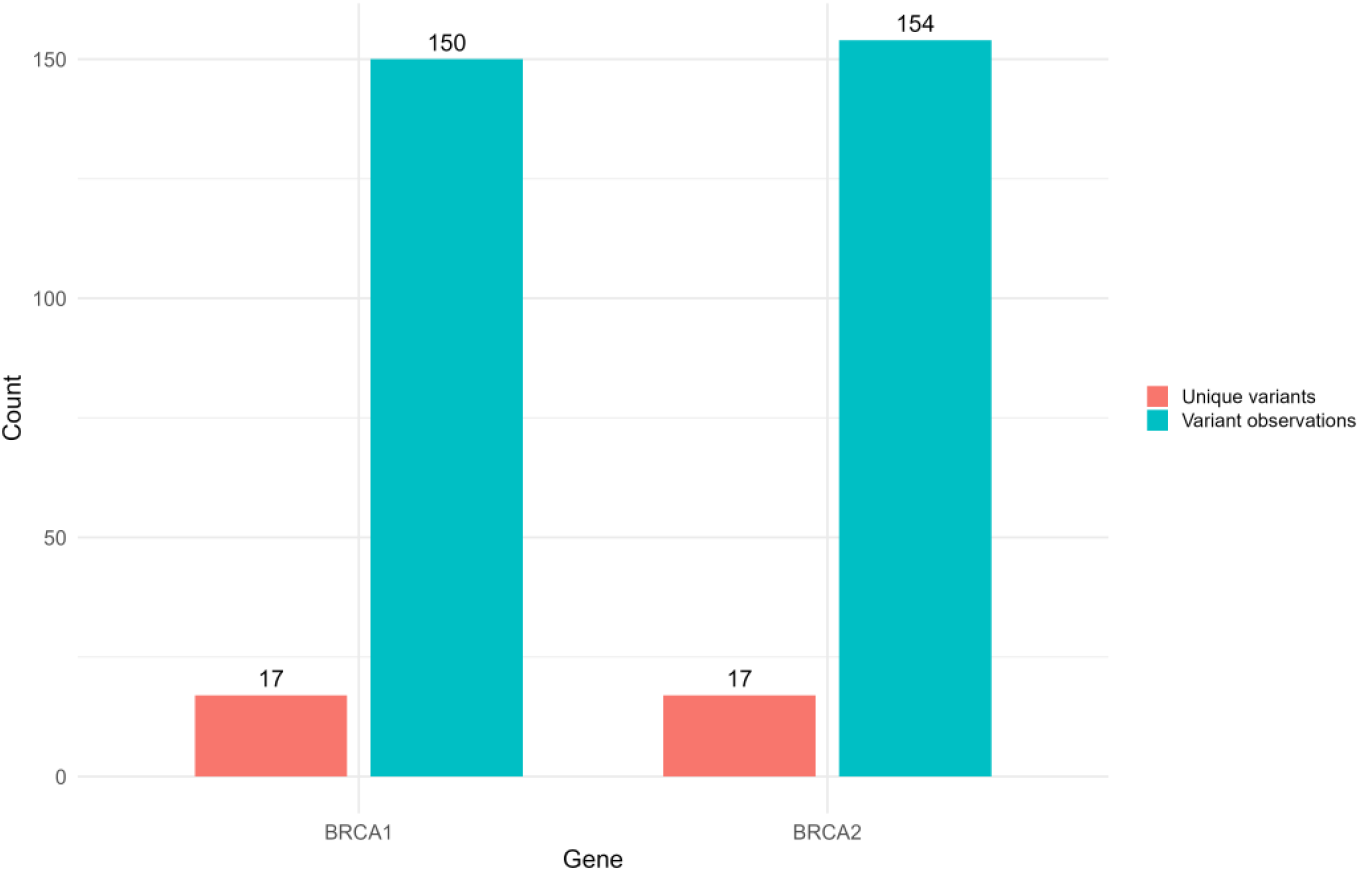
Variant observations and unique variants per gene. Bar plot comparing high-confidence Devyser-derived variant observations and unique variants in BRCA1 and BRCA2 across the analytical cohort. BRCA1 contributed 150 observations and 17 unique variants, while BRCA2 contributed 154 observations and 17 unique variants.

### Consequence profile and per-sample distribution

The main high-confidence analytical dataset was dominated by missense and synonymous variants. Across consequence annotations, missense variants were the largest class (141), followed closely by synonymous variants (135). Less frequent categories included intronic/splice-region annotations (15; 14 intron_variant and one splice_region_variant&intron_variant), 5′ UTR variants (7), frameshift variants (5), and one stop-gain variant. (**Figure 2**).

**Figure 2.**
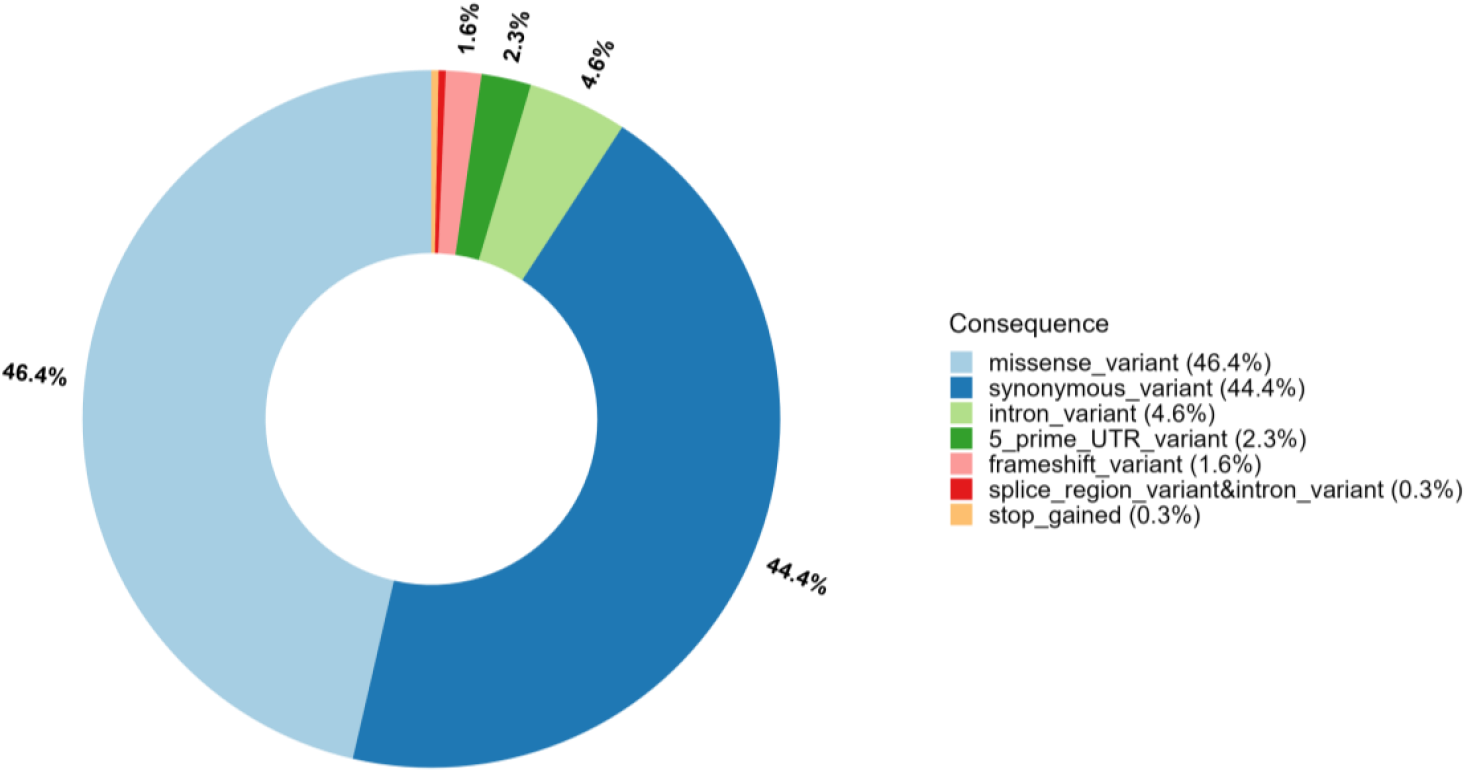
Distribution of variant consequence types. Overall distribution of Devyser-derived consequence annotations across the analytical cohort. Missense and synonymous annotations predominated; frameshift and stop-gain annotations were less frequent and sample-specific.

Per-sample consequence profiles showed broadly similar shared missense/synonymous polymorphic patterns, with frameshift or stop-gain consequences restricted to specific samples (**Figure 3**).

**Figure 3.**
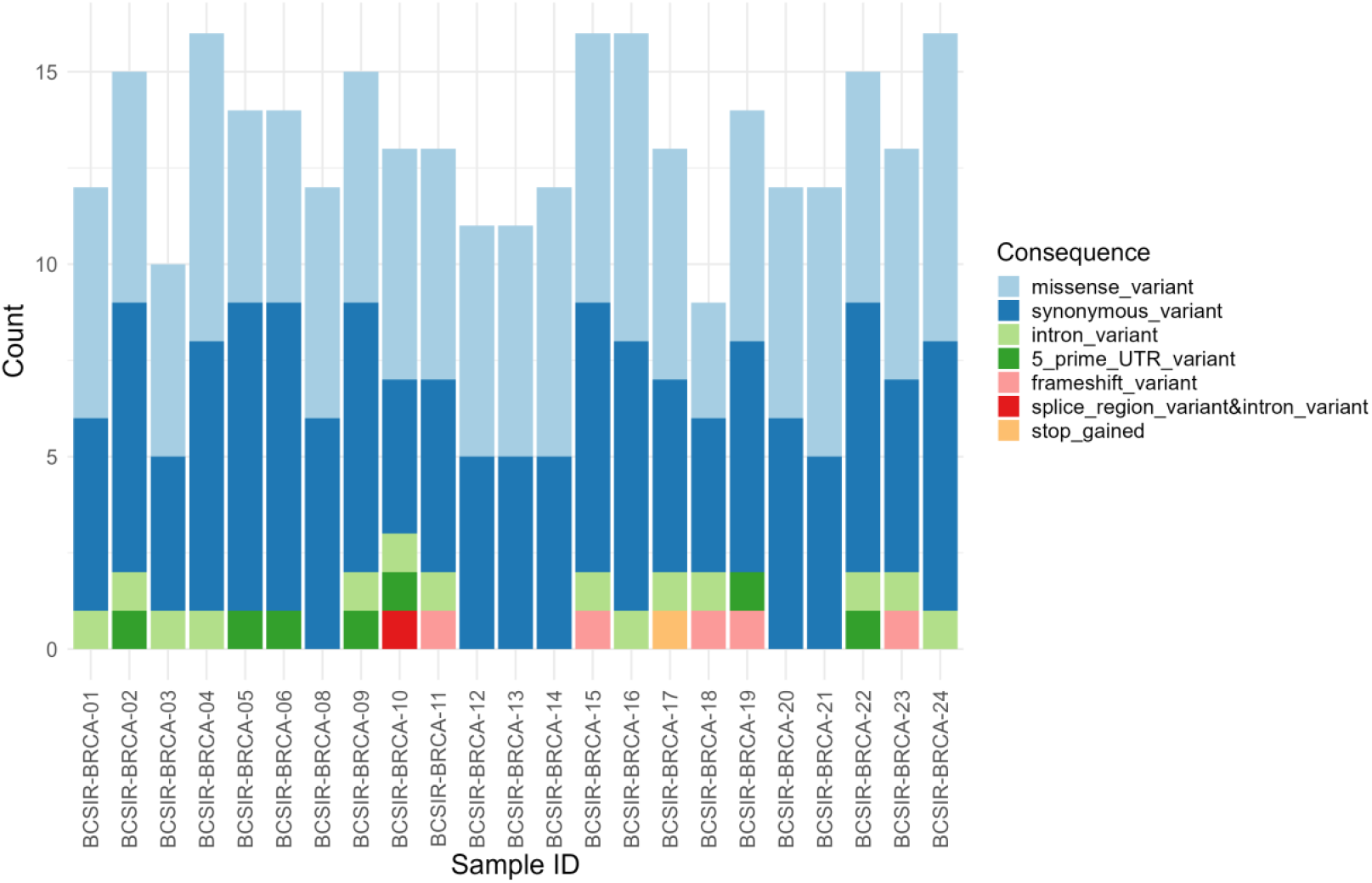
Consequence types per sample. Stacked bar plot showing the distribution of Devyser-derived consequence classes across individual samples. Most samples showed predominantly missense and synonymous variants, whereas frameshift and stop-gain consequences were restricted to specific samples.

The high proportion of synonymous variants was not interpreted as evidence of pathogenicity. Synonymous BRCA1/BRCA2 variants can occasionally influence splicing, but such effects require variant-specific evidence such as RNA analysis or validated functional data (Farber-Katz et al., 2018; Li et al., 2022). In this cohort, shared synonymous variants were therefore described as observed cohort-level signals rather than clinically significant hotspots.

### Candidate BRCA1 loss-of-function analytical findings

Six sample-specific heterozygous BRCA1 predicted loss-of-function short-variant candidates were identified (**Table 3**). These included five frameshift variants and one stop-gain variant. Variant allele fractions ranged from 45.47% to 50.81%, consistent with heterozygous germline-range observations, and total read depths ranged from 1159 to 6027. All were PASS/VCC-H calls in the Devyser output and were visually reviewed in IGV.

**Table 3.** Candidate BRCA1 loss-of-function analytical findings identified in the Devyser analytical callset.

| Sample | cDNA change | Protein change | Consequence | VAF (%) | Depth | Embedded Devyser ClinVar |
| --- | --- | --- | --- | --- | --- | --- |
| BCSIR-BRCA-11 | NM_007294.4:c.5038_5041del | p.Ile1680LeufsTer9 | Frameshift | 47.93 | 6027 | No embedded annotation |
| BCSIR-BRCA-15 | NM_007294.4:c.3756_3759del | p.Ser1253ArgfsTer10 | Frameshift | 47.13 | 1379 | Pathogenic |
| BCSIR-BRCA-17 | NM_007294.4:c.4508C>A | p.Ser1503Ter | Stop gained | 50.81 | 1159 | Pathogenic |
| BCSIR-BRCA-18 | NM_007294.4:c.4891dup | p.Ser1631LysfsTer48 | Frameshift | 45.47 | 1392 | Pathogenic |
| BCSIR-BRCA-19 | NM_007294.4:c.4065_4068del | p.Asn1355LysfsTer10 | Frameshift | 49.23 | 2216 | Pathogenic |
| BCSIR-BRCA-23 | NM_007294.4:c.2974del | p.Thr992LeufsTer8 | Frameshift | 48.26 | 2965 | No embedded annotation |

*Note: LoF, loss of function; VAF, variant allele fraction. Embedded ClinVar labels refer to the Devyser output database version ClinVar\_20190715 and are not current definitive clinical classifications. All listed variants are analytical loss-of-function candidates and require orthogonal confirmation and formal clinical curation before clinical reporting; the table does not represent final ACMG/AMP clinical classification.*

Four of the six candidate loss-of-function variants carried embedded ClinVar_20190715 pathogenic labels. Two frameshift variants, BRCA1 c.5038_5041del and BRCA1 c.2974del, did not have embedded ClinVar pathogenic labels in the assay output and were not straightforwardly resolved as pathogenic records during database cross-checking. They were retained in the candidate table because they were high-confidence heterozygous predicted frameshift calls with IGV support, but they are reported conservatively as analytical loss-of-function candidates requiring orthogonal confirmation and formal clinical curation.

### Protein-level and oncoprint variant patterns

Protein-level summaries showed shared BRCA1 and BRCA2 variants across the cohort (**Figures 4 and 5**). In BRCA1, commonly shared non-synonymous and synonymous signals included p.Pro871Leu, p.Lys1183Arg, p.Glu1038Gly, p.Ser1613Gly, p.Ser1436=, p.Leu771=, and p.Ser694=. In BRCA2, shared signals included p.Val2466Ala, p.Val2171=, p.Leu1521=, and p.Asn372His. These protein-level observations were interpreted as cohort-level common or polymorphic variant signals rather than pathogenic hotspots. In the BRCA1 indel/truncating panel, the candidate loss-of-function events were positioned between p.Thr992LeufsTer8 and p.Ile1680LeufsTer9, whereas no BRCA2 indel/truncating event met the main high-confidence candidate criteria.

**Figure 4.**
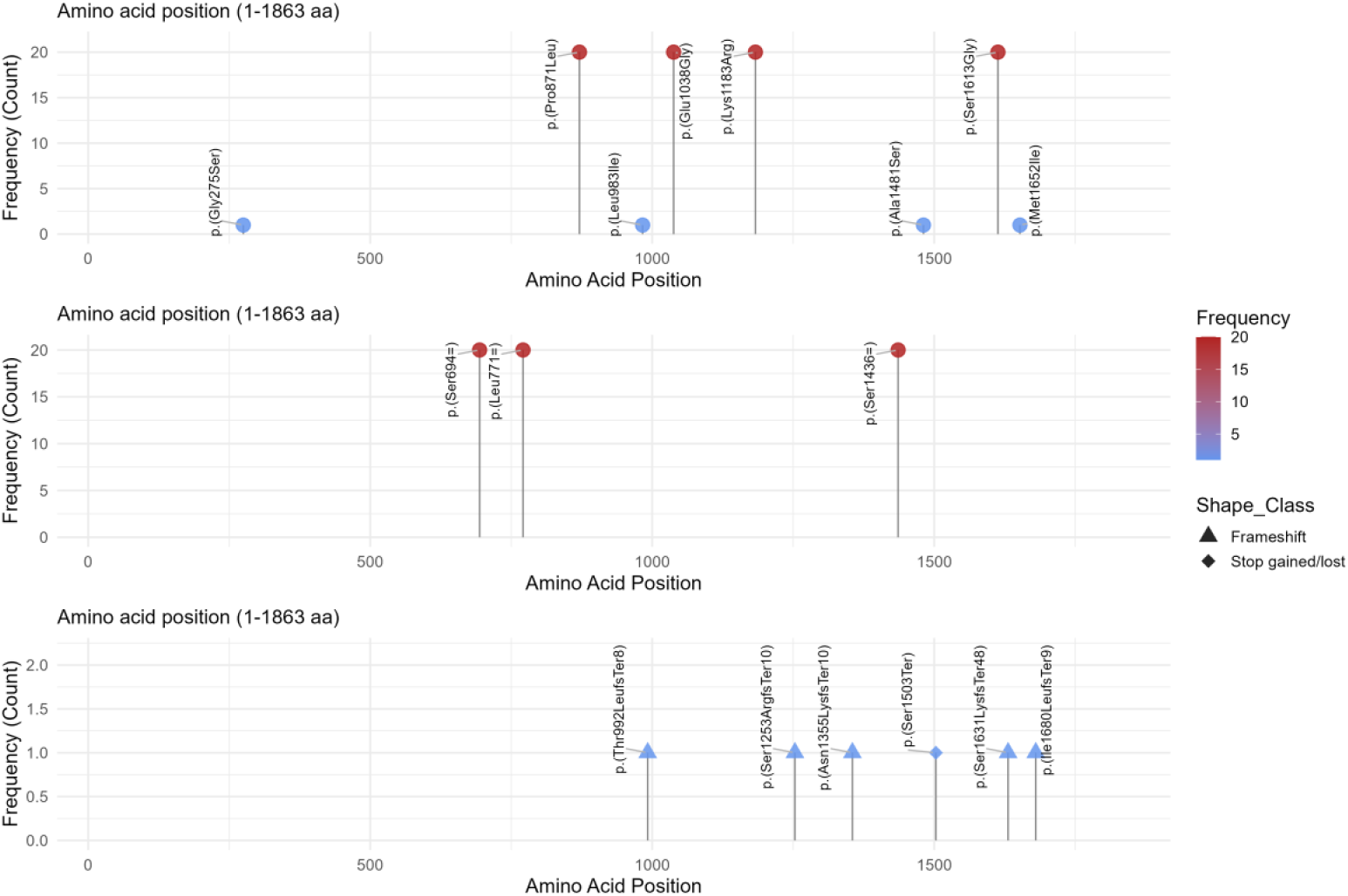
Protein-level variants in BRCA1. Three-panel lollipop plot showing BRCA1 non-synonymous substitutions, synonymous variants, and indel/truncating variants across the analytical cohort. Shared missense and synonymous variants are shown alongside sample-specific candidate loss-of-function events, which were positioned between p.Thr992LeufsTer8 and p.Ile1680LeufsTer9.

**Figure 5.**
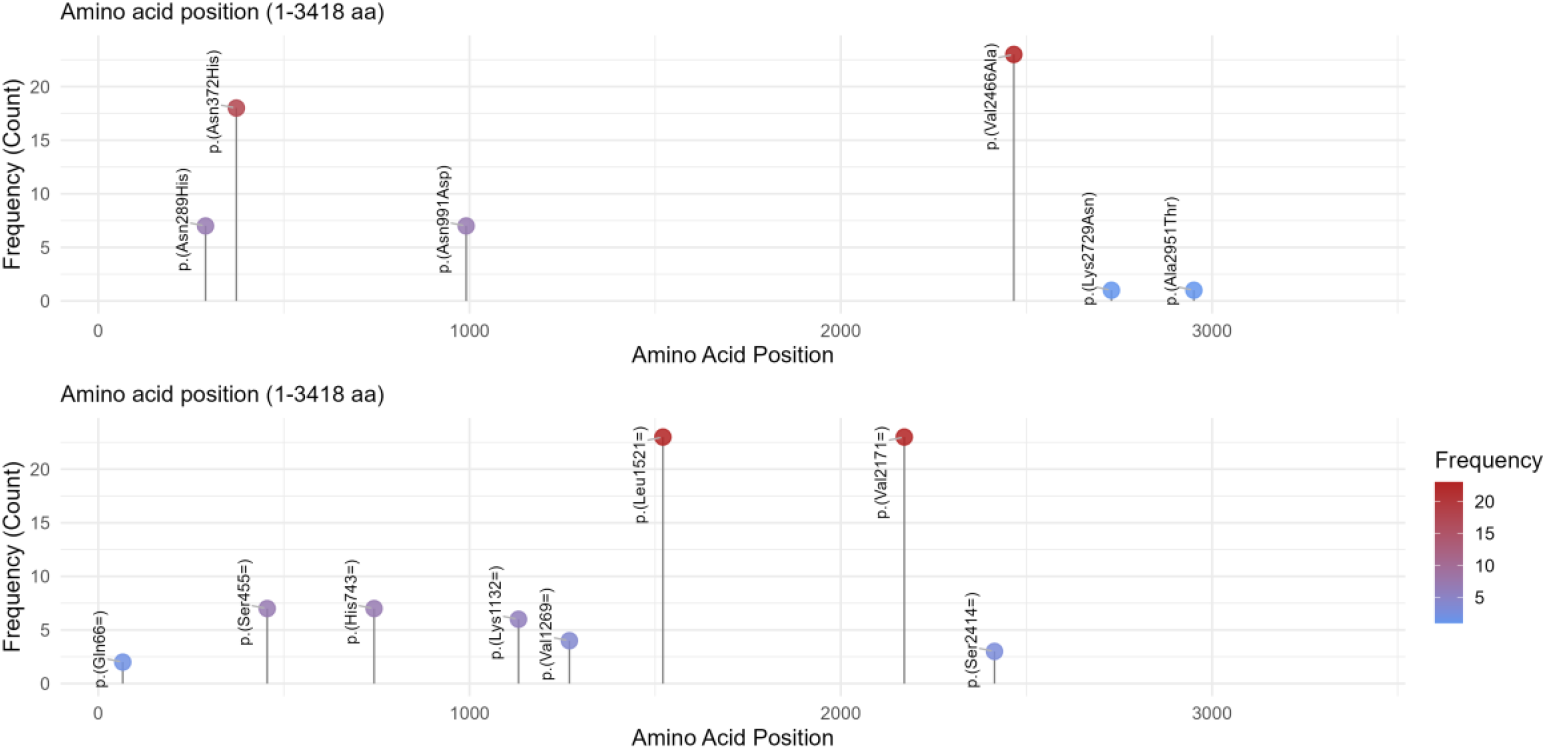
Protein-level variants in BRCA2. Three-panel lollipop plot showing BRCA2 non-synonymous substitutions, synonymous variants, and indel/truncating variants. BRCA2 signals were dominated by common missense and synonymous variants; no BRCA2 indel/truncating variant passed the main high-confidence candidate criteria.

The oncoprint-style heatmap further separated widely shared protein-level variants from sample-specific candidate loss-of-function events (**Figure 6**). The candidate BRCA1 frameshift and stop-gain variants appeared as isolated sample-specific events rather than cohort-wide truncating signals.

**Figure 6.**
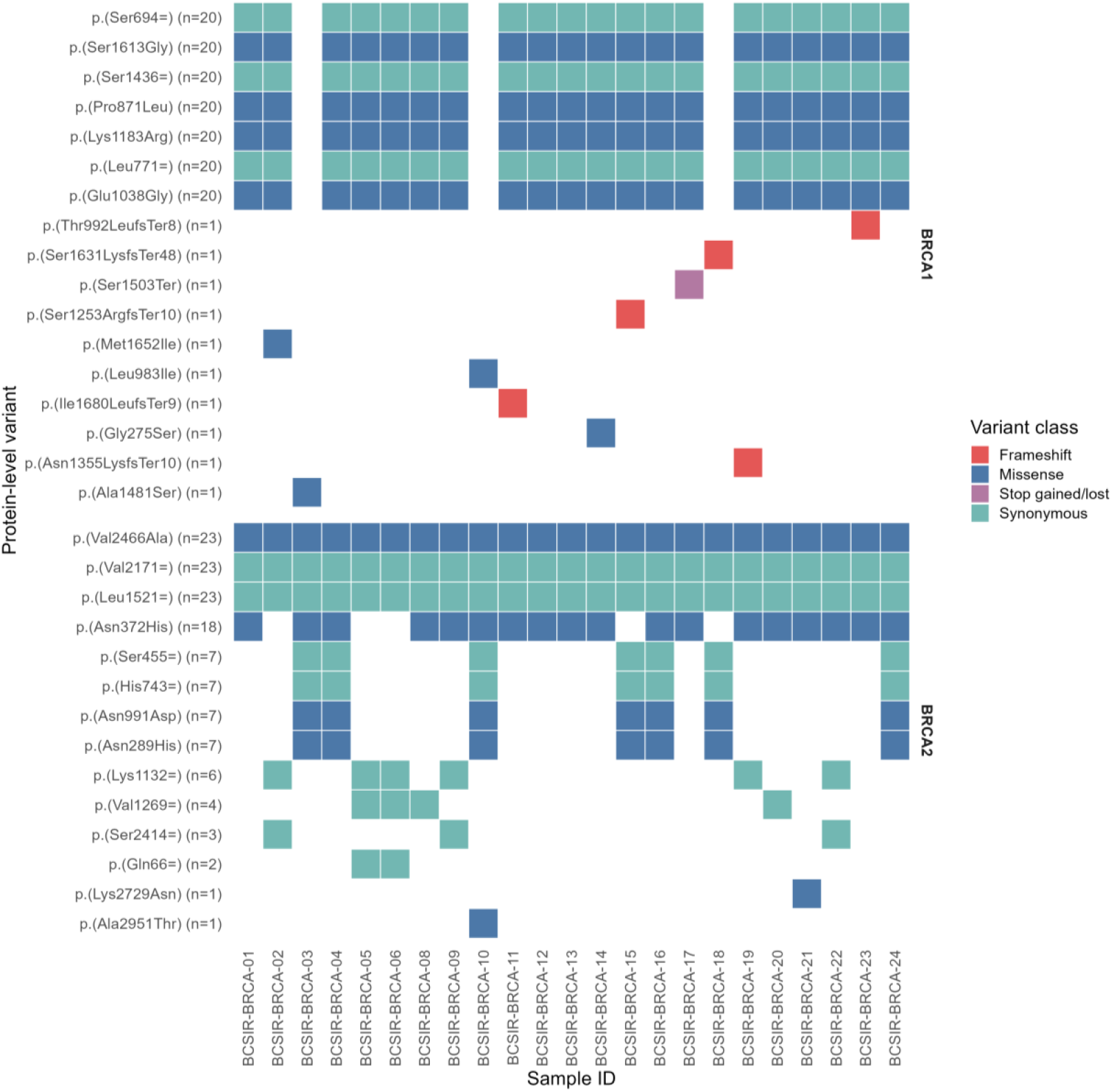
Oncoprint-style heatmap of observed protein-level BRCA variants. Heatmap showing the sample-wise distribution of observed protein-level variants across the analytical cohort. Widely shared variants were largely missense or synonymous, whereas candidate BRCA1 loss-of-function variants appeared as sample-specific frameshift or stop-gain events. This pattern supports separation of shared polymorphic signals from candidate analytically relevant loss-of-function findings.

### Embedded Devyser ClinVar annotation summary

Embedded Devyser ClinVar_20190715 annotations were summarized descriptively at the variant-observation level. Among 304 observations in the main analytical set, 207 were categorized as benign/likely benign, 89 as conflicting interpretations, 4 as pathogenic/likely pathogenic, and 4 had no embedded ClinVar annotation (**Figure 7**). These labels were not used as definitive clinical classifications.

**Figure 7.**
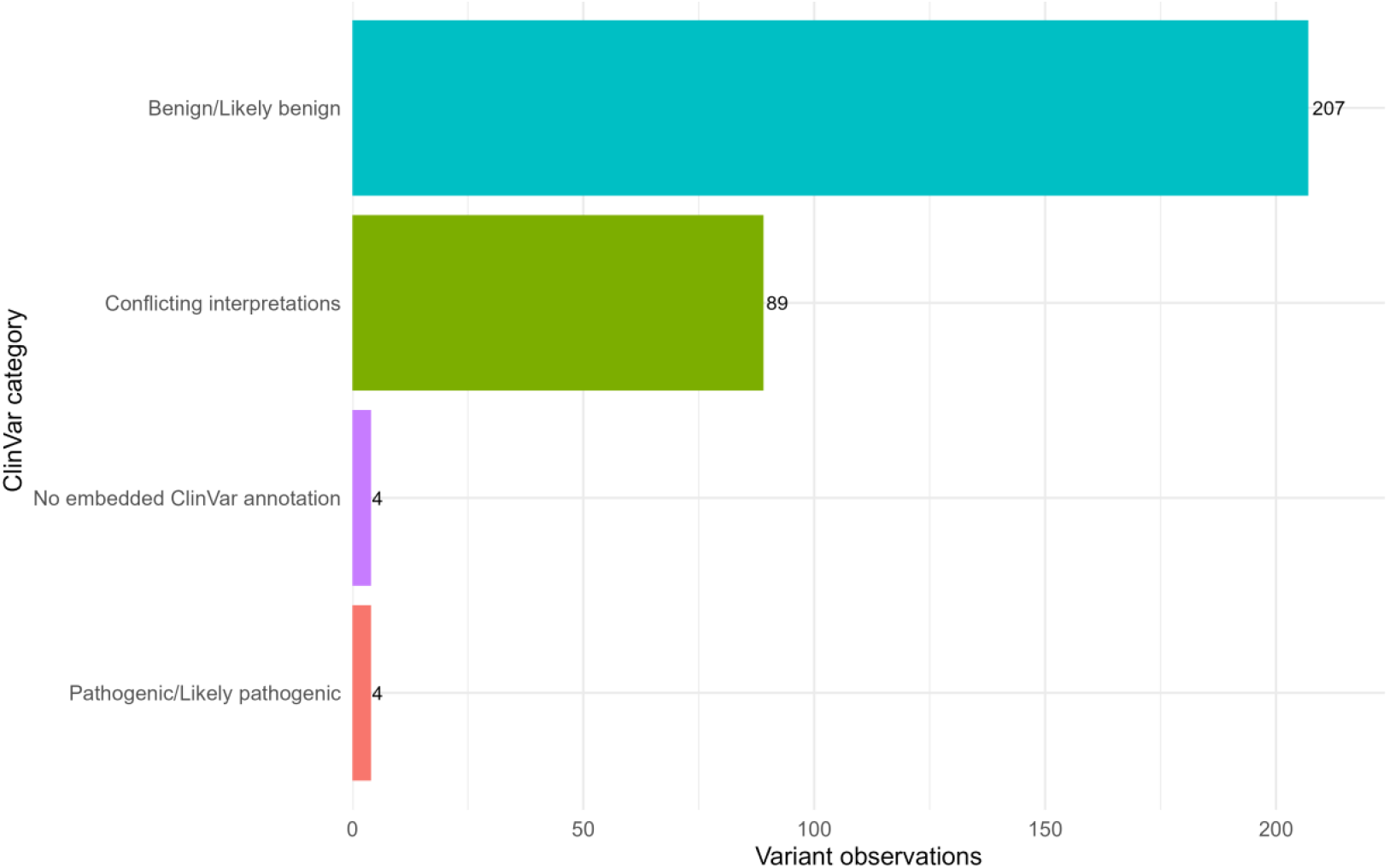
Embedded assay ClinVar annotation summary. Bar plot showing simplified ClinVar annotation categories extracted from the assay output. The output reported ClinVar_20190715; therefore, these annotations were summarized descriptively. Conflicting ClinVar interpretations were maintained as a separate category and were not interpreted as clinically actionable without manual curation.

### Candidate BRCA1 deletion-like CNV signal

Target-region and amplicon-level CNV outputs identified one notable candidate BRCA1 deletion-like CNV signal in BCSIR-BRCA-21 (**Figure 8**). Target-region CNV values were reduced across BRCA1 exons 15-20, clustering around approximately 0.50-0.54: exon 15, 0.5350; exon 16, 0.5112; exon 17, 0.5232; exon 18, 0.5047; exon 19, 0.5128; and exon 20, 0.5024. Flanking regions were near the diploid range, including exon 13 and exon 21.

**Figure 8.**
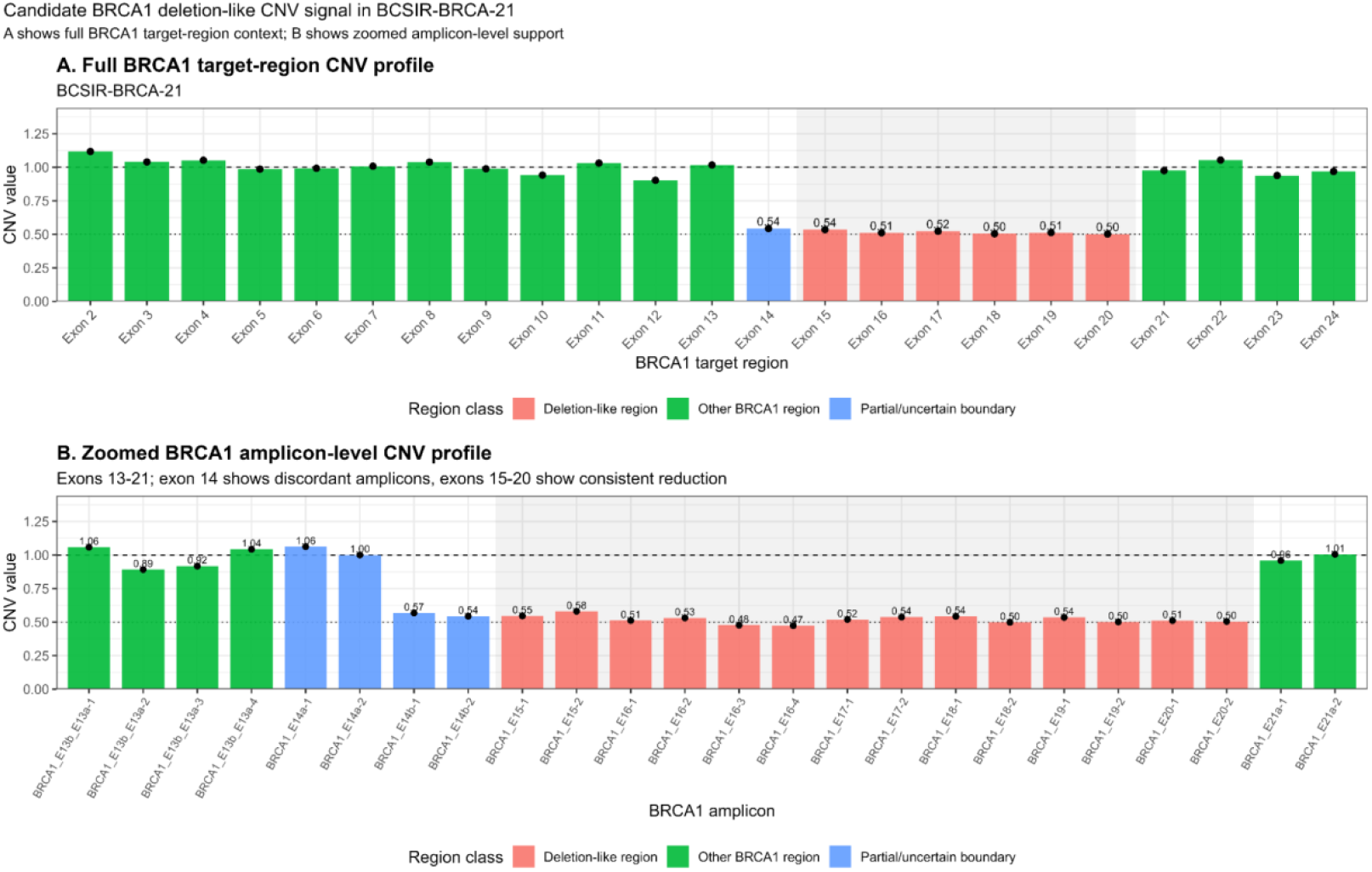
Candidate BRCA1 deletion-like CNV signal in BCSIR-BRCA-21. A, Full BRCA1 target-region CNV profile showing reduced copy-number values across BRCA1 exons 15-20, with flanking regions near the diploid range. B, Zoomed amplicon-level CNV profile across BRCA1 exons 13-21 showing concordant reduction across exons 15-20 and discordant exon 14 amplicon support, with BRCA1_E14b reduced and BRCA1_E14a near diploid range. The event was interpreted as a candidate BRCA1 exons 15-20 deletion-like CNV with possible/uncertain partial exon 14 involvement, requiring orthogonal confirmation.

Exon 14 showed discordant evidence. The target-region CNV value for BRCA1 exon 14 was reduced, but amplicon-level values showed that BRCA1_E14b amplicons were reduced while BRCA1_E14a amplicons remained near the diploid range. Specifically, BRCA1_E14b-1 and BRCA1_E14b-2 showed CNV values of approximately 0.57 and 0.54, whereas BRCA1_E14a-1 and BRCA1_E14a-2 were approximately 1.06 and 1.00. Therefore, the event was interpreted conservatively as a candidate BRCA1 exons 15-20 deletion-like signal with possible but uncertain partial exon 14 involvement. Because no orthogonal CNV confirmation was performed, this event requires confirmation by MLPA, ddPCR, or another validated CNV method before clinical reporting.

## Discussion

This study provides a conservative targeted BRCA1/BRCA2 analytical profile from a Bangladeshi clinically referred cohort. The main contribution is not estimation of BRCA1/BRCA2 variant prevalence, but demonstration of a transparent referral-cohort workflow that identifies candidate short-variant and CNV findings requiring confirmatory testing in a Bangladesh-relevant hereditary cancer context.

The primary analytical strength is that the manuscript is built around a single assay-specific short-variant, coverage, and CNV workflow, with explicit filtering and annotation-version reporting. This approach yielded a biologically plausible targeted BRCA profile dominated by missense and synonymous variants, with predicted loss-of-function events appearing as sample-specific findings.

The coverage-based QC summary further supports analytical adequacy of the assay outputs used in this study. All amplicon-sample measurements in the uploaded coverage table were at or above 100x and 200x, with only four measurements below 500x. These data support coverage adequacy for the descriptive targeted sequencing analysis. However, coverage depth alone does not replace full sequencing-run QC or orthogonal confirmation of candidate findings.

The clinical relevance of the study lies in the identification of a focused set of candidate BRCA1 predicted loss-of-function short variants and one candidate BRCA1 multi-exon deletion-like CNV signal. In tumor-suppressor genes such as BRCA1, frameshift and nonsense variants are biologically plausible loss-of-function candidates; however, in this study they remain analytical candidates because orthogonal confirmation and formal clinical classification were not performed (Richards et al., 2015).

Notably, the six candidate BRCA1 loss-of-function short variants mapped to the central-to-C-terminal coding portion of BRCA1, spanning p.Thr992LeufsTer8 to p.Ile1680LeufsTer9. This distribution is biologically plausible because truncating variants in this interval would be expected to remove or disrupt downstream BRCA1 regions involved in DNA-damage response and homologous recombination repair, including the serine cluster/SQ-cluster region, the PALB2-binding coiled-coil domain, and the BRCT-containing C-terminal region (Ismail et al., 2024; Roy et al., 2012). However, because the cohort was small and referral-enriched, this apparent regional concentration should not be interpreted as a Bangladeshi hotspot or population-level enrichment. Instead, it provides a protein-domain context for interpreting the candidate loss-of-function findings and prioritizing them for orthogonal confirmation and formal clinical curation.

The CNV finding is particularly important because short-variant-only analysis would miss exon-level deletions or duplications. The BCSIR-BRCA-21 signal showed consecutive reductions across exons 15 - 20 with supporting amplicon - level evidence, making it more compelling than an isolated target fluctuation. However, exon 14 involvement remains uncertain because E14a amplicons were near diploid while E14b amplicons were reduced. This is why the event is best described as a candidate exons 15 - 20 deletion - like CNV with possible partial exon 14 involvement, rather than a confirmed exon 14 - 20 deletion. From a testing - workflow perspective, this candidate CNV signal also illustrates why BRCA1/BRCA2 analysis in hereditary breast and ovarian cancer should include dosage - sensitive approaches rather than short - variant detection alone. Large genomic rearrangements, including exon-level deletions and duplications, represent an established class of BRCA1/BRCA2 pathogenic variation and may be missed by sequencing approaches that are not designed or validated for copy-number assessment (McVeigh et al., 2017). The consecutive reduction across BRCA1 exons 15-20 in BCSIR-BRCA-21 is therefore analytically noteworthy, but its exact breakpoint structure and exon 14 involvement cannot be resolved from the present assay output alone. Confirmation using MLPA, ddPCR, or another validated CNV method would be required before this event could be considered reportable or clinically interpretable.

The country and regional context is also important. Bangladesh remains underrepresented in hereditary breast and ovarian cancer genomics, and previous local studies have mainly assessed selected BRCA1 exons or smaller variant sets (Chowdhury et al., 2020; Nishat et al., 2019). South Asian reviews show substantial heterogeneity in BRCA1/BRCA2 findings across populations, indicating that founder or shared patterns reported in one country should not be assumed to apply uniformly across the region (Kharel et al., 2022). The present cohort is not a prevalence study, but it contributes a carefully filtered local pilot dataset and identifies candidate variants for confirmatory follow-up.

The expanded referral metadata provide useful context for the candidate analytical findings. Most samples with candidate BRCA1 loss-of-function short variants had documented positive family history, and the sample with the candidate BRCA1 multi-exon deletion-like CNV also had positive family-history documentation. Nevertheless, the metadata were incomplete, non-uniformly recorded, and not collected under standardized hereditary cancer testing criteria. These observations therefore should be interpreted as referral-context descriptors rather than genotype–phenotype associations.

The high burden of synonymous and shared polymorphic variants should be interpreted cautiously. Synonymous variants are often benign or likely benign, but selected synonymous BRCA1/BRCA2 changes can affect splicing through effects on exon-intron recognition or exonic splicing regulatory elements (Farber-Katz et al., 2018; Li et al., 2022). In the current study, no RNA-level or functional assays were performed; therefore, shared synonymous observations were not interpreted as clinically significant without supporting evidence.

Embedded ClinVar annotation was dominated by benign/likely benign and conflicting interpretation categories. This pattern is expected when embedded ClinVar fields are summarized at the observation level and emphasizes why parsed database labels should not be treated as final clinical classifications. Candidate loss-of-function variants were separately cross-checked against public database records during manuscript preparation where possible, and the results are provided in **Supplementary Data File 1**. These checks support contextual interpretation but do not replace ACMG/AMP-aligned manual curation or orthogonal laboratory confirmation (Landrum et al., 2018; Richards et al., 2015). Formal ACMG/AMP or ENIGMA-aligned clinical classification was outside the scope of this analytical pilot study; therefore, all candidate variants are presented as laboratory-derived candidates requiring confirmatory testing and expert clinical curation.

This study has several limitations. First, the cohort was small and clinically referred, so frequencies should not be interpreted as general Bangladeshi population prevalence. Second, clinical metadata were incomplete, limiting genotype-phenotype correlation. Third, orthogonal confirmation was not performed for candidate short variants or the CNV signal. Fourth, the embedded annotation database was historical, and current clinical classification requires updated manual curation. Fifth, while IGV review strengthened read-level confidence, it is not a substitute for Sanger confirmation of short variants or MLPA/ddPCR confirmation of CNVs. Sixth, the coverage table supports targeted depth adequacy but does not provide a complete independent sequencing-run QC profile.

Despite these limitations, the study provides a focused and defensible starting point for Bangladesh-relevant BRCA interpretation. Future studies should include larger phenotype-linked cohorts, updated database annotation, orthogonal validation, systematic CNV confirmation, ACMG/AMP- or ENIGMA-aligned curation, and family-based assessment of segregation.

## Conclusions

Targeted BRCA1/BRCA2 analysis using an assay-specific AmpliconSuite workflow on an Illumina NextSeq 550 platform identified a high-confidence short-variant profile in a Bangladeshi clinically referred cohort. The coverage profile supported analytical adequacy of the targeted sequencing outputs, with all amplicon-sample measurements at or above 100x. The callset was dominated by shared missense and synonymous polymorphic variants, while six sample-specific candidate BRCA1 predicted loss-of-function short variants and one candidate BRCA1 exons 15-20 deletion-like CNV signal were identified. All clinically relevant candidates require orthogonal confirmation and formal clinical curation before clinical reporting. The findings provide a conservative pilot dataset for future validated Bangladesh-specific hereditary breast and ovarian cancer genomics research.

## Data Availability

De-identified clinical metadata, assay-derived short-variant tables, CNV tables, candidate variant tables, public database cross-checks, and coverage-based QC summaries are provided in the supplementary data files. Additional de-identified supporting data may be shared upon reasonable request, subject to institutional and ethical approval requirements.

## Supplementary Information

**Supplementary Data File 1** contains the Devyser-only supplementary workbook, including sample-level QC summary, high-confidence short-variant observations, all PASS/VCC-H short variants, unique variants, candidate BRCA1 loss-of-function variants, public database cross-checks for candidate loss-of-function variants, low-VAF review variants, target-region CNV values, amplicon-level CNV values, CNV review events, figure index, and data dictionary.

**Supplementary Data File 2** contains the raw amplicon-level coverage table and per-sample coverage-based QC summary used for the coverage analysis in this manuscript.

**Supplementary Data File 3** contains de-identified and generalized clinical referral metadata, including non-overlapping 5-year age bands where age information was available, referral indication category, generalized family-history status, TNBC documentation status, candidate-finding context. Precise ages, specific family relationships, and raw clinical notes were removed or generalized for participant privacy. These metadata were incomplete and retrospectively curated from referral/laboratory intake documentation; therefore, they were used only for descriptive cohort characterization.

## Declarations

### Consent for publication

All participants provided consent for anonymized publication of study findings.

### Availability of data and materials

De-identified and generalized clinical metadata, assay-derived short-variant tables, CNV tables, candidate variant tables, public database cross-checks, and coverage-based QC summaries are provided in the supplementary data files. The embedded annotation database versions reported in the assay output were ClinVar_20190715 and dbSNP_b151. Additional de-identified supporting data may be shared upon reasonable request, subject to institutional and ethical approval requirements.

### Authors’ contributions

Syed Muktadir Al Sium: conceptualization, writing - original draft, writing - review and editing, software, data curation, formal analysis, visualization, investigation, methodology. Tanjina Akhtar Banu: writing - review and editing, conceptualization, data curation, formal analysis, investigation. Barna Goswami: writing - review and editing, data curation. Showti Raheel Naser: investigation, writing - review and editing. Md Ahashan Habib: resources. Shahina Akter: writing - review and editing. Mst. Hosne Ara: resources. Sheikh Md Selim Al Din: resources. Ali Nafisa: resources. Maksudur Rahman Nayem: resources. Mohammad Fazle Alam Rabbi: resources. Md Murshed Hasan Sarkar: conceptualization, resources, supervision, project administration, methodology, writing - original draft, writing - review and editing, visualization, software. Md Salim Khan: supervision, project administration, resources, conceptualization.

## Acknowledgements

The authors sincerely thank Sanjana Fatema Chowdhury, Scientific Officer, BCSIR, for her valuable review and feedback. The authors also acknowledge the staff of Invent Technologies Ltd. for their technical support during sequencing. The authors are grateful to all participants who voluntarily provided samples for this study. The authors further thank Abdur Rahman and all others who assisted with blood collection.

